# Performance of A Point-of-care Fluorescence Immunoassay Test to measure anti Severe Acute Respiratory Syndrome Corona Virus 2 Spike, Receptor Binding Domain Antibody Level

**DOI:** 10.1101/2023.11.01.23297911

**Authors:** Marita Restie Tiara, Hofiya Djauhari, Febi Ramdhani Rachman, Antonius Christianus Rettob, Darmastuti Utami, Fahda Cintia Suci Pulungan, Heru Purwanta, Rudi Wisaksana, Bachti Alisjahbana, Agnes Rengga Indrati

## Abstract

Quantitative determination of anti-SARSCoV2-S-RBD is necessary for the evaluation of vaccination effectiveness. Surrogate viral neutralization test (SVNT) is approved for measuring anti-SARSCoV2-S-RBD, but a point-of-care platform is needed to simplify anti-SARS-CoV-2-S-RBD measurement. We aimed to evaluate the performance of a rapid fluorescent immunoassay-based kit, FastBio-RBD^tm^, compared to SVNT. During April–September 2021, we enrolled two groups of subjects; convalescent subject and subject without COVID-19 history. Subjects were tested for anti-SARS-CoV2-S-RBD antibody using FastBio-RBD^tm^ and GenScript-cPASS^tm^ SVNT. We measured the correlation coefficient and conducted ROC analysis to determine the best cut-off value of anti-SARSCoV2-S-RBD against SVNT percent inhibition levels of 30% and 60%. We included 109 subjects. Anti-SARS-CoV-2-S-RBD strongly correlated to SVNT %-inhibition with R value = 0,866 (p < 0,0001). ROC analysis showed that anti-SARS-CoV-2-S-RBD of 6,71 AU/mL had 95,7% sensitivity and 87,5% specificity to detect percentage inhibition of 30%. Anti-SARS-CoV-2-S-RBD of 59,76 AU/mL had a sensitivity of 88,1% and specificity of 97,0% to detect percentage inhibition of 60%. FastBio-RBD^tm^ can determine the presence and level of anti-SARS-CoV-2-S-RBD with good sensitivity and specificity. It has the potential to be deployed in health facilities with limited resources.

## Introduction

Coronavirus disease-19 (COVID-19), the infection of Severe cute Respiratory Syndrome Coronavirus-2 (SARS-CoV-2) has become a pandemic.[1] COVID-19 vaccination and monoclonal antibodies have been one of the main strategies to face this problem. COVID-19 vaccination programs aim to reduce severe disease and death from COVID-19.[2] A myriad of COVID-19 vaccines have been developed and used on different platforms, with different efficacy and immunogenicity.[3] Comorbidities in a person may impact immunogenicity evoked by vaccines; some comorbidities may impair antibody response to vaccines.[4] An individual’s immune system response is the most important marker for determining mortality and morbidity.[5] In the early COVID-19 pandemic, there was limited scientific evidence on the development and duration of the immune response in individuals, both in cases of infection and post-vaccination against SARS-CoV-2.[6] Antibody testing after vaccination can help identify the people who are low responders to vaccines and personalize the use of vaccines in these people. A person with a negative or low antibody level early in the disease can benefit from the provision of monoclonal antibody.

Virus-neutralizing antibodies confer protection by blocking the interaction that mediates virus entry into susceptible host cells. For SARS-CoV-2, this interaction involves binding of the receptor binding domain (RBD) of the SARS-CoV-2 spike glycoprotein with the angiotensin-converting enzyme 2 (ACE2).[7] The gold standard test of immune response and level of protection after SARS-CoV-2 vaccination is the plaque-reduction neutralization test (PRNT). However, PRNT is tedious and can only be done in sophisticated laboratories with biosafety level (BSL)-3.[8] A surrogate-viral neutralization test (SVNT) produced by GenScript was approved as sensitive and specific to detect neutralizing antibody in 2021.[9] The Genscript SARS-CoV-2 SVNT is a commercially available assay that detects antibody that specifically inhibits the RBD-ACE2 interaction without using live SARS-CoV-2.[7] However, this test is based on competitive enzyme-linked immunoassay reaction, hence the need to properly handle equipment in a standardized immunological laboratory.[8] Given the width and breadth of the pandemic, it is imperative to have point-of-care testing, which can be implemented practically and safely in simple laboratories to identify people who are low-responders to vaccines.

Lateral flow immunoassay is a simple technology that allows point-of-care testing to be done and has already been used in numerous settings (public health centres, hospitals, home-care-based test) for several biomarker targets such as the detection of human chorionic gonadotropin (hCG) in pregnancy test kits, Human Immunodeficiency Virus (HIV) tests, and others.[10] It is also affordable and can be implemented in the laboratory with limited resources, allowing for massive application. However, they can only provide qualitative results, hence unsuitable for antibody testing. Fluorescence immunoassay technology can make the application of lateral flow assays quantifiable with the simplicity of lateral flow assays.[11] Similarly, this latter technology has been used in various tests like determination of inflammatory markers like CRP, PCT, and HBa1C for diabetes screening.[12]

The objective of this study is to evaluate the performance of a point-of-care, fluorescence immunoassay kit that measures the level of antibody to SARS-CoV-2: FastBio-RBD, compared to a surrogate-viral neutralization test.

## Materials and Methods

### Study Design

We conducted a cross-sectional survey enrolling subjects from April until September 2021. Subjects were invited based on their voluntary willingness and stratified based on a sample block of convalescent subjects (with a history of COVID-19) and subjects without a history of COVID-19. Vaccinated subjects were examined around 2.5 - 4 months and between 4 – 6 months after full dose vaccination. We chose vaccinated subjects who had been vaccinated for two and a half months after vaccination because we would like to ensure that the anti-S-RBD results measured were predominantly IgG, without much influence from IgA and IgM. This had been known to occur within 2.5 months after completion of vaccination.[13] All subjects consented to participate in the study and have no known uncontrolled comorbidity.

### Data collected

We collected data on demographics, such as age, sex, history of COVID-19 illness, vaccination status, type of vaccine, and vaccination interval. We collected data on comorbidities, such as diabetes, hypertension, chronic pulmonary diseases, and chronic kidney diseases.

### Ethical Clearance

The study was approved by the Health Research Ethics Committee of Dr. Hasan Sadikin General Hospital, Universitas Padjadjaran with ethics number 410/UN6.KEP/EC/2021, 17^th^ May 2021. The study was conducted in accordance with the Declaration of Helsinki, and all data were kept anonymous.

### Reference test: GenScript cPass SARS-CoV-2 Neutralization Antibody Detection Kit

The GenScript cPass SARS-CoV-2 Neutralization Antibody Detection Kit (Genscript Biotech, Leiden, Netherlands), SVNT, was used as the reference test. The test was performed according to the manufacturer’s instructions.[14] Briefly, serum samples, as well as negative and positive controls were diluted 1:10 in sample dilution buffer, mixed 1:1 with HRP-RBD working solution and incubated at 37 °C for 30 min. Subsequently, 100 μL of sample and controls were added into the wells of the 96-well plate, which have been coated with the ACE2 receptor protein. The plate was incubated at 37 °C for 15 min and washed 4×with 300 μL of washing buffer. Next, 100 μL of substrate solution was added and the plate was incubated in the dark for 15 min at RT. Finally, 50 μL of stop solution per well was added and the absorption at 450 nm was measured using an ELISA-Reader. The percentage of signal inhibition in relation to the negative control was calculated as follows:

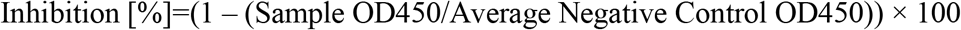

### Test being validated: FastBio-RBD—SARS-CoV-2 Antibody test

The FastBio-RBD test was produced by Wondfo (Wondfo Biotech Co.Ltd Guangzhou, China) for distribution in Indonesia by PT Biofarma Indonesia (persero), FastBio-RBD. The test was performed according to the manufacturer’s instructions.[15] It was based on a point-of-care SARS-CoV-2 RBD Antibody test using fluorescence immunoassay technology. It was an immunochromatographic test using an upregulated phosphorylation indicator. The platform was based on a sandwich reaction where the test line, contained S-RBD antigen. Serum samples were added to the detection buffer, mixed, and added to the sample well. The patient’s anti-S-RBD antibodies in the serum would bind with the RBD antigen conjugated with a phosphorescent marker and form immune complexes. The immune complexes then migrate on the nitrocellulose membrane, which is then captured by the RBD antigen in the test line. The resulting complex would be detected by the related Fluorescent Immuno-Assay (FIA) Meter. Quantification was enabled by the fluorescence intensity exited from the immunochromatographic test. Anti-S-RBD result was then displayed as Arbitrary Unit (AU) /mL. The higher the antibody in the sample, the more fluorescent the complex, and therefore the higher the results of anti-S-RBD. According to the manufacturer, 20 times multiplication will convert FastBio-RBD AU result to Binding Antibody Unit (BAU), which the World Health Organization recommended. BAU is the first WHO International Standard for anti-SARS-CoV-2 immunoglobulin.

### Statistical Analysis

Demographic and baseline characteristics were presented in a frequency tabulation. Numeric values of the test result were presented with a median and IQR. The anti-S-RBD results measured with FastBio-RBD were reported in AU/mL. The percent inhibition measured with SVNT was reported in %.

The correlation between the FastBio-RBD result and the % inhibition result of the SVNT test was determined by Spearman’s ranked test after the log transformation of the values. Based on the value of both tests, we also determined the ROC for the detection of specific levels of inhibition. Next, based on the ROC, we determined the sensitivity and specificity of FastBio-RBD at % inhibition cut-off of 30%, which was the positive cut-off given by the SVNT kit[14] and cut-off of 60%, which had been determined to correlate with a vaccine efficacy of 70 – 90%.[16] All calculations were done using IBM SPSS version 25 (IBM Corporation, New York, United States of America). Graphics were further refined using GraphPad Prism version 8.0. (Graphpad Software, LLC, San Diego, United States of America).

## Results

### Study sample

We enrolled 109 subjects in this study, 58 of which were post-COVID-19 subjects in the convalescent phase and 51 without COVID-19 history. Baseline characteristics are shown in Table 1. Thirty (51.7%) subjects of the convalescent group had received two vaccination doses. Fifteen (29.4%) subjects among those without a history of COVID-19 were never vaccinated with any COVID-19 vaccines, while 36 (70.6%) had completed two doses of vaccinations. Besides the vaccination status proportion, there are no statistically significant characteristic differences between the two groups. All subjects in both groups who had been vaccinated received CoronaVac, an inactivated virus vaccine.

**Table 1.** Baseline Characteristics of the Participating Subjects.

| Characteristics | Without History of COVID-19<br>(N = 51) | Convalescent<br>subjects<br>(N = 58) |
| --- | --- | --- |
| Sex, n (%) |  |  |
| Male | 24 (47,1) | 31 (53,4) |
| Female | 27 (52,9) | 27 (46,6) |
| Age, (years) |  |  |
| 18 – 29 | 15 (29,4) | 14 (24,1) |
| 30 – 39 | 15 (29,4) | 17 (29,3) |
| 40 – 49 | 8 (15,7) | 13 (22,4) |
| 50 – 59 | 3 (5,9) | 6 (10,3) |
| ≥60 | 10 (19,6) | 8 (13,8) |
| Comorbidity, n (%) |  |  |
| No comorbidity | 39 (76,4) | 43 (74,1) |
| Yes | 12 (23,6) | 15 (25,9) |
| Hypertension | 8 (15,7) | 3 (5,2) |
| Diabetes mellitus | 1 (2,0) | 4 (6,9) |
| Cardiovascular disease | 2 (3,9) | 2 (3,4) |
| Chronic lung disease | 0 (0) | 5 (8,6) |
| Others | 1 (2,0) | 1 (1,7) |
| **Among subjects without History of COVID, n (%)** |  |  |
| No vaccination | 15 (29,4) | 28 (48,3) |
| Vaccinated | 36 (70,6) | 30 (51,7) |
| Time interval between COVID-19 vaccination and testing n (%) |  |  |
| 2.5 – 4 months | 18 (35,3%) | n.a. |
| ≥4 months | 18 (35,3%) | n.a. |
| **Among subject with with history of COVID-19** |  |  |
| Time interval between test and COVID-19, n (%) |  |  |
| <1 month | n.a. | 14 (24,1) |
| 1 – 2 months | n.a. | 26 (44,8) |
| 2 – 3 months | n.a. | 8 (13,8) |
| >3 months | n.a. | 10 (17,2) |

\* n.a. : not applicable

### Anti-S-RBD titer of convalescent subjects and vaccinees without COVID-19 history measured using FastBio-RBD

The distribution of anti-S-RBD antibody levels of all subjects is shown in Figure 1a. Among the unvaccinated subjects, we saw four persons who already had high antibody titers. Two persons were also shown to have high antibody titers in the group who had been vaccinated. We observed slightly higher anti-S-RBD titer in subjects without COVID-19 history at 2.5 – 4 months compared to the group with > 4 months after vaccination. However, this increase was not statistically significant. The anti-S-RBD titers of the convalescent subjects were significantly higher than the rest of the groups.

**Figure 1a.**
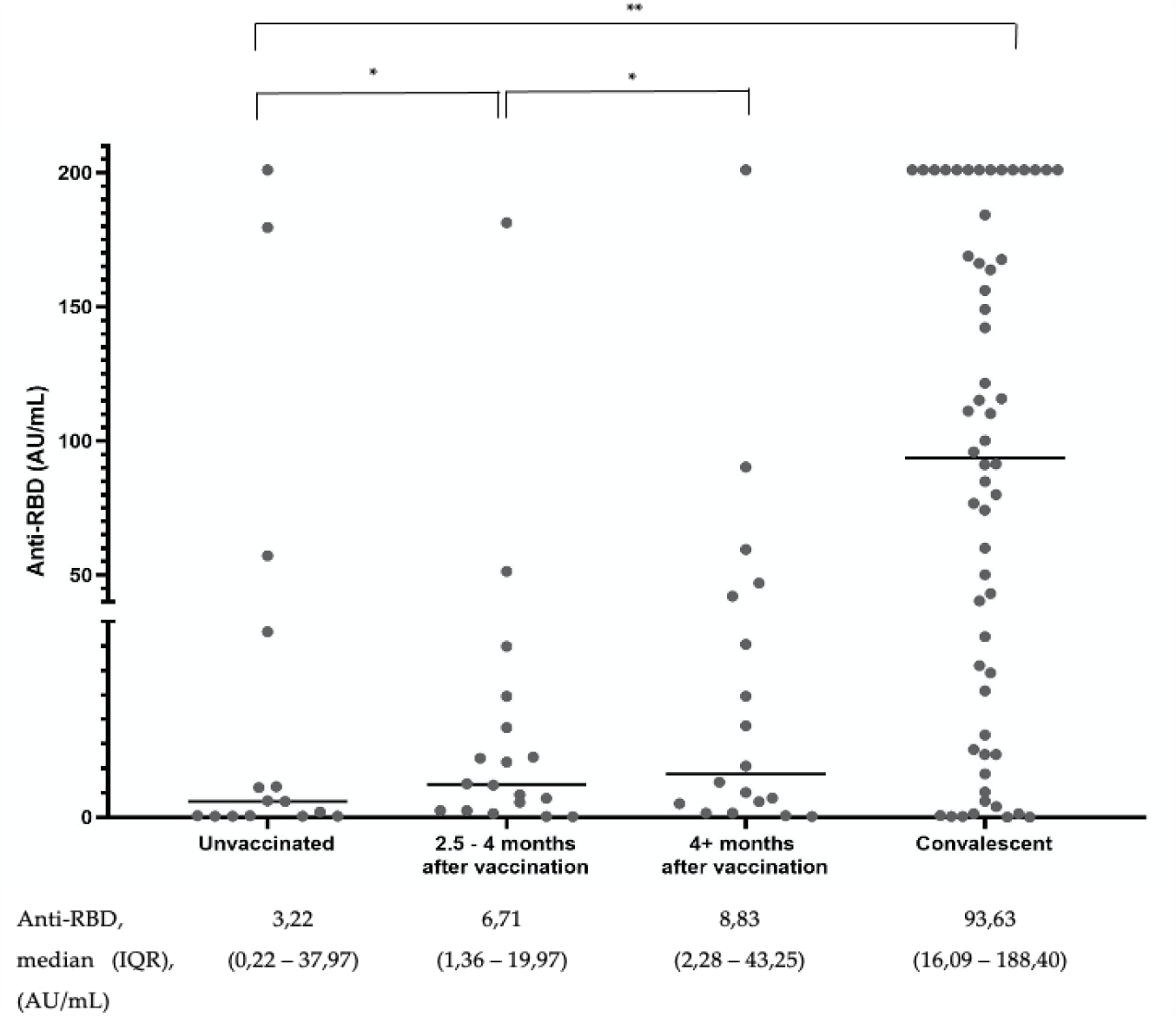
Distribution of anti-S-RBD antibody level among all subjects. The graph represents a plot of the result, and the horizontal line depicts the median value. *\*p value* ≥ *0.05, **p-value < 0.05*

### Percentage inhibition of convalescent subjects and vaccinees without COVID-19 history measured using SVNT

The distribution of SVNT % inhibition of all subjects measured using the SVNT test is shown in Figure 1b. Similarly, % inhibition in subjects without COVID-19 history at 2.5 – 4 months and > 4 months after vaccination was shown to be statistically higher compared to the unvaccinated group. We also see that the % inhibition among convalescent subjects showed statistically significantly higher values than the rest of the group. SVNT test was conducted to be more sensitive than anti-S-RBD to detect the presence of inhibition. From the same subject serum, we observed that 51 subjects (46.7%) have reached higher than mid to high range value of SVNT, whereas only 33 subjects (30.2%) in the anti S-RBD tests.

**Figure 1b.**
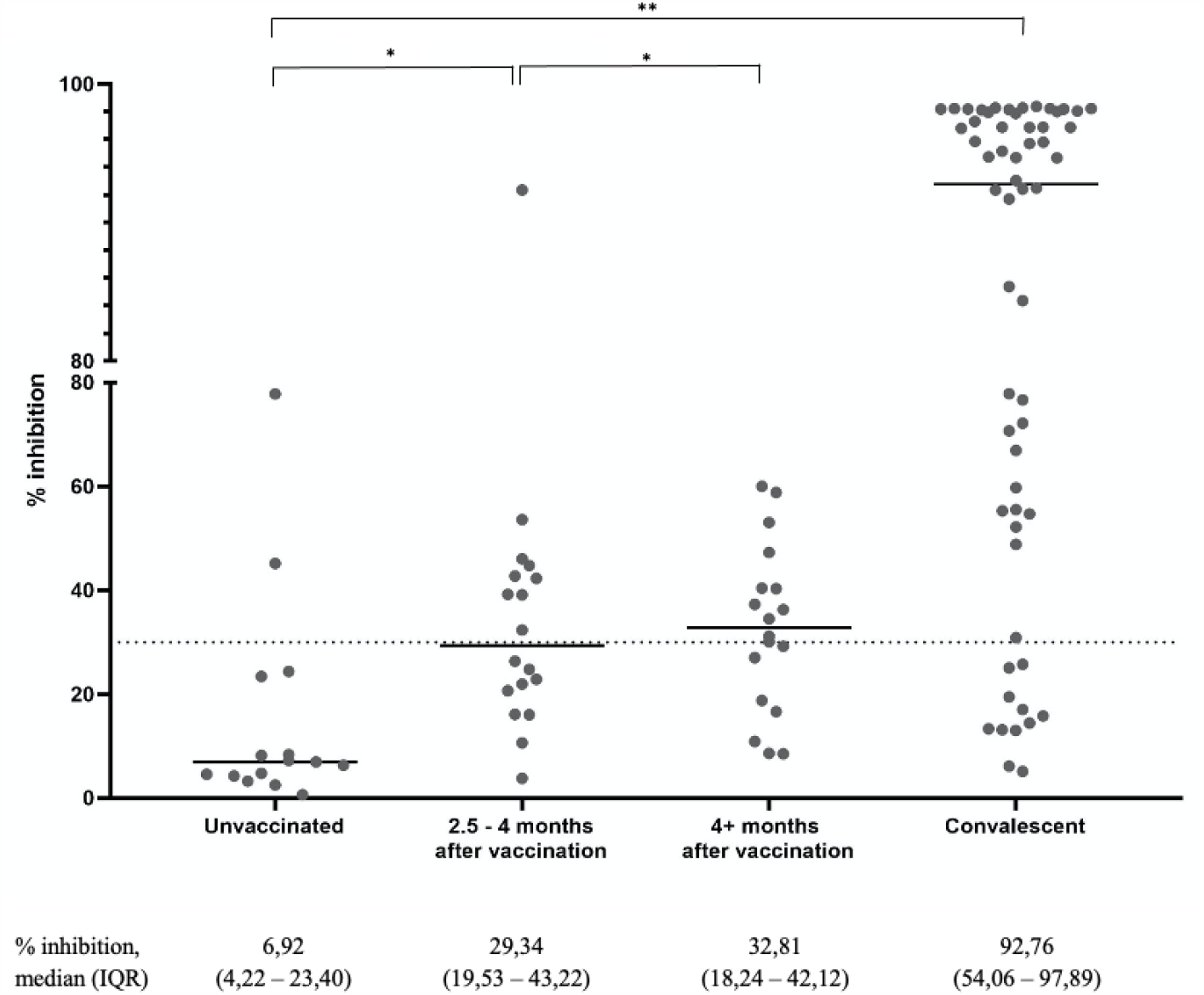
Distribution of Surrogate Viral Neutralization percent inhibition among all subjects. The graph represents a plot of the result. Horizontal lines depict the median value. *\*p value* ≥ *0.05, **p-value < 0.05*

### Correlation between anti-S-RBD titer and percentage inhibition

The Spearman’s ranked correlation between anti-S-RBD titer and the % inhibition also had very strong correlation, with R = 0,866 (95% CI 0,808, 0,908), p < 0,0001 (Figure 2).

**Figure 2.**
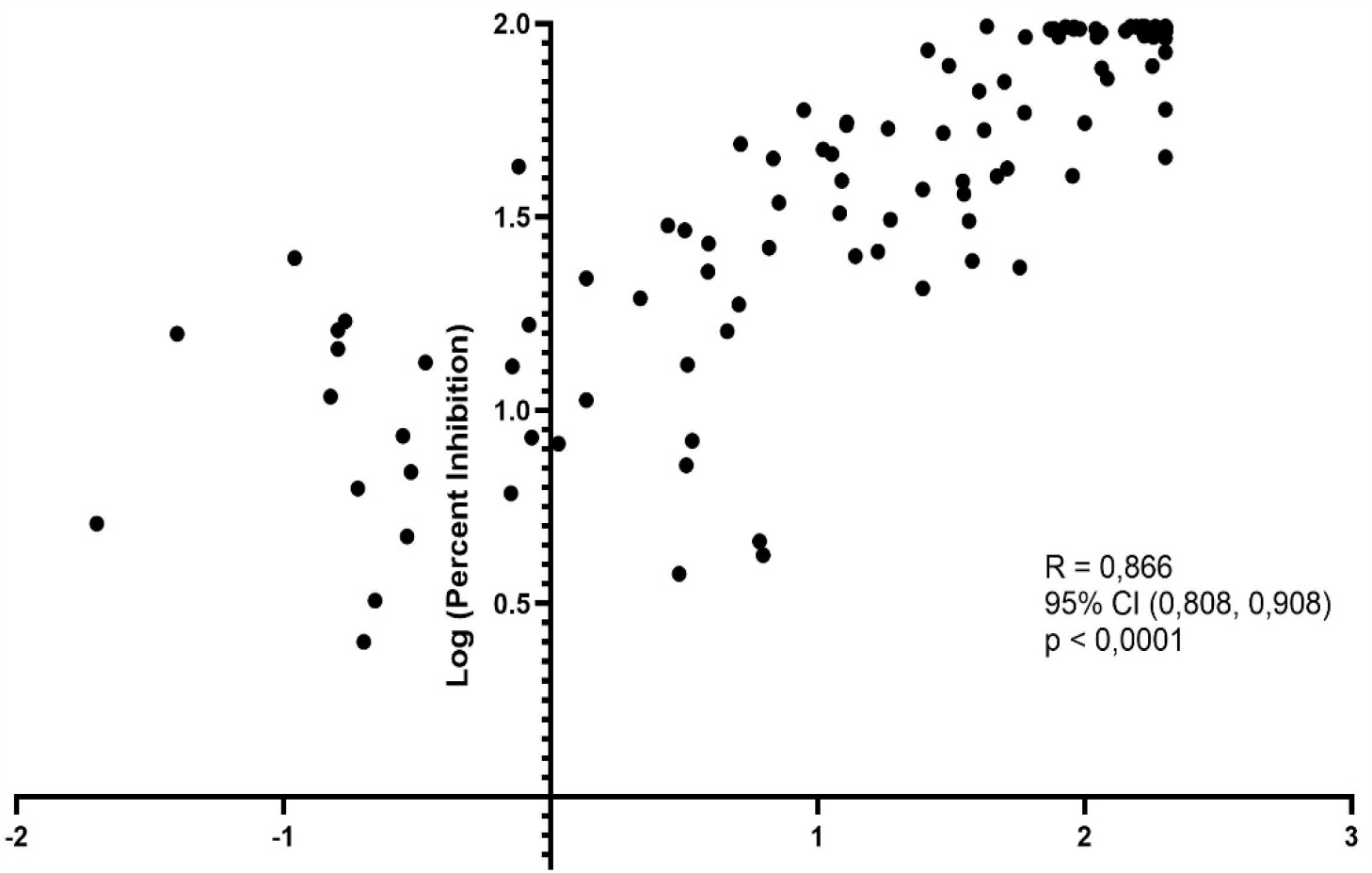
Spearman’s correlation between anti-S-RBD antibody level to Surrogate Viral Neutralization test % inhibition among all subjects

### Accuracy of FastBio-RBD vs SVNT

We performed three ROC analyses to determine the best cutoff of the FastBio-RBD kit using the 30% and 60% inhibition by SVNT as the standard. The first ROC curve corresponded to 30% inhibition showing an excellent AUC of 0,957, 95% CI (0,922, 0,992), p < 0.0001. Using this curve, we obtained an anti-S-RBD value of 6,71 AU/mL (134,2 BAU/mL) as the best cut-off. With this cut-off value, the accuracy of FastBio-RBD to detect 30% inhibition was very good (Table 2).

**Table 2.**
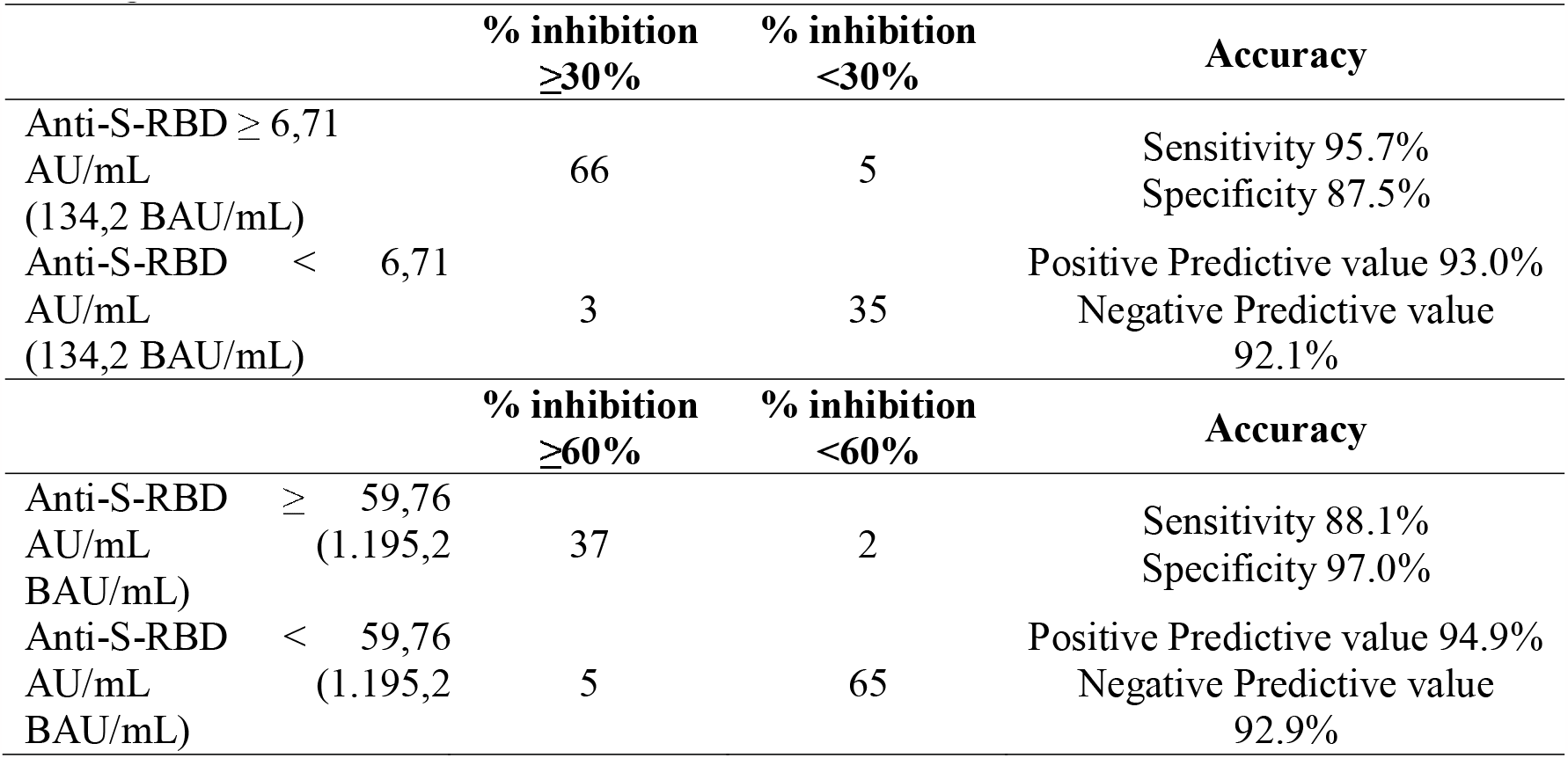
Accuracy of anti-S-RBD at bet cut-off point for 30% and 60% inhibition level of Surrogate Viral Neutralization.

Next, we plotted the ROC curve using the cut-off inhibition percentage of 60% which also showed an excellent AUC 0,956, 95% CI (0,917, 0,995), p <0,0001. Using this curve, we obtained the best cutoff point for an anti-S-RBD value of 59,76 AU/mL (1.195,2 BAU/mL). With this value, the FastBio-RBD detected 60% inhibition with positive predictive value and negative predictive value of 94.9% and 92.9% respectively (Table 2).

## Discussion

This study shows that FastBio-RBD shows adequate accuracy in detecting presence of antibody to SARS-CoV-2 with good agreement to GenScript SVNT. We have shown that the parameters properly show the increasing antibody level among the vaccinated individuals. The results correlate well with GenScript SVNT results across wide range of values. And with the proper cut-offs, we can show good test accuracy to detect protection levels of 30% and 60%.Quantitative detection of SARS-CoV-2 NAb titres is crucial to screen therapeutic antibodies from convalescent patients, predict humoral protection, evaluate vaccine efficacy, and optimize immunization strategies.

GenScript SVNT is a good surrogate test to PRNT and can be applied as a screening test to detect the presence of neutralization antibodies against SARS CoV-2 if a test is needed.[9] Due to the lack of availability of PRNT in our country, we use the Genscript SVNT as the gold standard for evaluating the FastBio-RBD.

There are many commercial serological assays based on the principles of enzyme immunoassays (EIA), fluorescence immunoassays (FIA), and chemiluminescent immunoassays (CLIA) have been developed to improve large-scale immunity test.[17] FastBio-RBD with its chromatographic fluorescence immunoassay technology has the potential to allow a faster detection and measurement of anti-S-RBD antibody in serum samples and can be used in point-of-care-testing settings. This practicality aspect is important in Indonesia, where a lot of the population lives in hard-to-reach areas where technically demanding laboratory procedures cannot be performed. In addition, as a standardized commercial test, the test results can be compared across multiple settings and easily converted to the standard unit of BAU/mL. This contrasts with tests like PRNT, where the antibody measured depends on the experimental setup (such as the type of virus particles used, dilution, and type of cell culture) and hence can only be compared with caution.

Anti-S-RBD and percentage inhibition measurements in our subjects showed that the anti-S-RBD is higher in convalescent subjects than in vaccinated subjects. This validation study was conducted in the second year of the pandemic, before the delta virus outbreak in Indonesia. Therefore, the overall neutralizing antibody level in the population was still low.[18] This is similar to the studies conducted in this period by Gluck et al in Germany.[19] In contrast to the mRNA vaccine, the inactivated virus vaccine does give less antibody response than the convalescent person.[20] The FastBio-RBD is sensitive enough to detect seropositivity, which can be shown to have positive (≥1 AU, or ≥20 BAU) among the vaccinated person. However, SVNT seemed to be more sensitive to detecting the presence of neutralization antibody than FastBio-RBD.

The correlation analysis showed that FastBio-RBD showed a very strong positive correlation to Genscript SVNT, comparable with more sophisticated tests like the ECLIA and ELISA-based test.[21] This result is also better in comparison with other immunochromatographic FIA tests. [21] Hence, the correlation coefficient provided in this paper can be considered for using this test to detect antibody response in SARS-CoV2.

The detection of the protection level is the main objective of the parameter. Using the suggested cutoffs at at 30% and 60% inhibition level, the FastbioRBD shows good results. The strength of the FastbioRBD is the high specificity or high positive predictive value (94.5%) to detect 60%. With this result, we can interpret that if a person has reached 59.8 AU/ml, there is a 97% probability that he has already reached a 60% protection level. A high negative predictive value to detect 60% inhibition means that if they have not reached 59.8% AU/ml, there is a 98% probability that this level has not been reached. This interpretation is necessary for deciding if a person needs additional booster. SVNT levels of 60% have been shown to correspond with 70% protection efficacy. [16]

Therefore, measurement of anti-S-RBD using FastBio-RBD in both convalescent and vaccinated subjects can detect accurately whether the anti-S-RBD measured meant sufficient protection against COVID-19. Individuals with adequate protection could be offered additional vaccinations, especially if the individuals had known immunocompromised states. Even more widely, the FastBio-RBD could also determine whether herd immunity has been achieved in a given area or community.[18] Here we resume the advantages and disadvantages of FastBio-RBD^tm^ and GenScript-cPASS^tm^ in table 3.[22,23]

**Table 3.** Advantages and Disadvantages of FastBio-RBD^tm^ (Fluorescence immunoassay) and GenScript-cPASS^tm^ (SVNT)

|  | Advantages | Disadvantages |
| --- | --- | --- |
| FastBio-RBD™<br>(Fluorescence immunoassay) | - Affordable and easily utilized in small clinical laboratories or point-of-care testing<br>- Test using various specimens, including finger-prick or veinpuncture whole blood, serum, or plasma.<br>- Cost-effective<br>- Comparable accuracy, good specificity especially at 60% inhibition<br>- Allows quantitative detection of antibodies. | - It cannot distinguish binding antibodies (bAbs) from neutralizing antibodies (nAbs)<br>- Less sensitivity at low neutralization antibody level |
| GenScript-cPASS™<br>(SVNT) | - Scalable alternative in comparison to conventional neutralization assays.<br>- More sensitive in detecting neutralizing antibodies (nAbs)<br>- Acceptable standard for various clinical and research settings. | - Need to be conducted in well equipped immunology lab.<br>- Elisa reading require skilled laboratory technician |

The limitation of our study was that we compared our anti-S-RBD results with GenScript SVNT only. Ideally, we should have compared the anti-S-RBD against PRNT. However, PRNT is currently unable to be performed in Indonesia. By comparing the anti-S-RBD to GenScript SVNT, which was a surrogate of PRNT, we can provide an approximation that the FastBio-RBD is indeed accurate.

## Conclusion

FastBio-RBD can detect immune response to COVID-19 or vaccine as a proxy of the neutralization antibody level they have acquired. It is a specific test that if we have reached an anti-RBD of >59.76 AU/ml, we can be certain at 94.9% that the person already has achieved a 60% inhibition level, corresponding to 70% vaccine efficacy. On the other hand, if he has not reached this level of anti-RBD, we can be 92.9% certain that he has not reached this level of inhibition. FastBio-RBD is based on the immunochromatographic FIA test and can be used at the point of care test, hence, it can be widely implemented even in the primary care setting.

## Data Availability

All data produced in the present work are contained in the manuscript

